# Genomic Epidemiology and Emerging Mechanisms of Antibiotic Resistance Among Clinically Significant Bacteria

**DOI:** 10.64898/2026.02.17.26346381

**Authors:** Ali J. Muhialdin, Ali M. Hussein, Rahman K. Faraj

## Abstract

**Background:** The never-ending emergence of superbugs casts a shadow over the victorious age of antibiotics. In fact, the triumph of antibiotics was previously viewed in retrospection as our final victory over bacteria. Bacteria like *Klebsiella pneumoniae, Acinetobacter baumannii*, and *Escherichia coli* are now raising an alarming number of infections across hospitals and communities around the globe. The objective was to evaluate the implications for antimicrobial stewardship based on identifying the antibiotic resistance profiles, genotype mechanisms, and trends in common pathogenic bacteria found in various hospitals across Iraq.

**Methods:** We used a two-fold approach that was comprehensive in scope and involved both efficient multicenter surveillance as well as cutting edge genetic analysis to unravel the complex topography of antibiotic resistance. We provided a geographically heterogeneous but diverse set of clinically obtained isolates to participate in hospitals for a period of 24 months and concentrated our efforts on prioritized pathogens *K. pneumoniae, A. baumannii, E. coli, P. aeruginosa*, and *S. aureus* that are well known to pose serious threats. Beginning with clinically obtained isolates sourced across the entire globe, we used standardized techniques such as broth microdilution to first undertake phenotyping in a central reference lab to establish microbial identity based on resistance phenotypes to a set of prioritized antibiotics that include carbapenems, third generation cephalosporins, or fluoroquinolones. Finally, we derived data concerning the emergence patterns and geographic distribution of resistant microbes such as MRSA or CRE. We used genome-wide sequencing to unlock information concerning the genetic blueprints for a set of specifically chosen isolates based on their representational diversity across geographic locales, resistance phenotypes, and specific times.

**Results:** The sample was made up of *Escherichia coli* (n = 225), *Klebsiella pneumoniae* (n = 185), *Staphylococcus aureus* (n = 135), *Pseudomonas aeruginosa* (n= 90), and *Acinetobacter baumannii* (n = 125). Ceftriaxone resistance was found in 80.4% of *E. Coli*, ciprofloxacin resistance in 45.6%, and meropenem resistance in 15.1%. *K. pneumoniae* exhibited 38.9% resistance to aminoglycosides and 70.2% resistance to carbapenems. The percentage of MRSA in S. aureus was 55.5%. P. aeruginosa showed 22.2% resistance to colistin, 37.8% resistance to piperacillin tazobactam, and 50.0% resistance to ceftazidime. Imipenem resistance was found in 85.6% of *A. baumannii* isolates, whereas colistin resistance was found in 28.8% of isolates. In all, 3.4% of isolates are pan-drug-resistant (PDR), 14.6% are extensively drug-resistant (XDR), and 52.1% are multidrug-resistant (MDR). WGS identified common genes such bla_NDM-1, bla_OXA-48, mcr-1, aac (6’)-Ib, and plasmid replicons IncF, IncL/M, and IncI2. Carbapenem resistance in Gram-negative bacteria rose by around 18% over the course of five years.

**Conclusions:** This study shows that the rapid spread of complex genetic information in bacteria causes antibiotic resistance problems. High-level resistance represents an expected consequence of the spread of resistance genes and successful bacteria within healthcare systems. We demonstrate in our results that our expertise in overcoming resistance at a molecular level will play a crucial role in combating infectious diseases in the coming years.

## Introduction

Imagine a scenario where the ‘wonder drugs’ used in the last 20th century are not effective. In other words, a ‘normal procedure’ carries a ‘fatal’ consequence. On top of that, a ‘minor scratch’ results in permanent infection. In fact, that’s almost the developing scenario concerning ‘the antibiotic resistance problem’—a so-called ‘silent pandemia’ which ‘shapes contemporary medicine’ to date. We used to ‘have an irrefutable edge’ in our ‘fight against microbes because of antibiotics (1).

These medications have been used for over a half-century to treat everything from run-of-the-mill strep throat to potentially life-threatening bouts of sepsis. But this golden age is coming to a close. Under the pressure of selection and driving forces of evolution, bacteria are fighting back. Our best hope against them is less effective with each passing generation because these microbes are not just surviving our efforts but learning as well (2).

But how real this new scenario actually is can well be understood against the background of the results that our multicenter monitoring project has obtained. We are no longer facing scattered resistance but are confronted with serious endemic threats. The statistics shown above form a patient record not only because there are numbers involved but because we are living in our contemporary healthcare environment. Indeed, nowadays resistance to carbapenems, which are a family of antibiotics to which *Acinetobacter baumannii* or *Klebsiella pneumoniae* are quite frequently resistant when serving as a last resort (3).

These “superbugs” are between 70-90% resistant to treatment in some areas. In fact, related to resistance to antibiotics, resistance to first line medications such as fluoroquinolones or third generation cephalosporins has skyrocketed to 70% in a common bacterium named *Escherichia coli*. In fact, *E. coli* causes urinary tract infections. In other words, if these percentages are true, those medications currently being used in doctor’s offices around the globe will not function (4).

The pressing environment found in hospitals, especially ICUs found within them, contributes to this challenge. In these settings, there are too many antibiotics used in a short period of time to highly susceptible patients. These settings create an environment that fosters the development and spread of resistant bacteria. The annex to our dataset illustrates how complex this problem is. The emergence or spread of resistant bacteria silently occurs due to international travel and patient migration. In other settings, there are pharmacies carrying antibiotics to patients directly without instructions prescribed by physicians. Apart from these challenges, there are catastrophic situations like displacement due to natural disasters or political unrest that are an extraordinary burden to these settings. In these settings, there are high instances of diseases emerging from bacteria that are classified as extensively-drug resistant (XDR)/pan-drug resistant (PDR), which are incurable because antibiotics are used. We face a ‘post-antibiotic era’ not in some distant tomorrow but today (5).

“How” in our response carries so much significance, but we are informed “what” and “where” there is resistance based on data derived from surveillance studies like ours. These trends that are apparent on the surface, but a reflection of a great biochemical battle based on genetic mutations. Carbapenem resistance poses a problem due to varying enzymes presented by these pathogens. These enzymes are NDM and OXA-48. These enzymes are actually molecular scissors that deftly cut drugs away before these pathogens are harmed.

In a similar fashion, Extended Spectrum Beta Lactamases (ESBLs), a resistance mechanism in *E. coli*, makes a significant contribution to its resistance to cephalosporins. Also, Metallo-beta-lactamase (MBL) producers pose a highly important risk to *P. aeruginosa*. In fact, owing to these resistance mechanisms being plasmid-encoded or acting as molecular shuttles which move easily across species into various species of bacteria, resistance to antibiotics correlates to unprecedented increases (6).

As a result, genetic analysis becomes a crucial subsequent procedure. The genetic design involved in the resistance exhibited by a bacterium belongs to the field of genomics. On analyzing traditional surveillance, we establish that our organism merely resists. We are able to detect its resistance genes (the “resistome”) to which our pathogens belong and its history in hospitals based on its DNA. The goal of our research titled “Emerging Trends and Mechanisms of Antibiotic Resistance in Clinically Important Pathogenic Bacteria” lies in bridging that gap. We are bringing our minds together to move past merely presenting data to understanding genetic tales (7).

In an effort to realize numerous highly important objectives using both surveillance analysis and cutting-edge genomic analysis, to first determine whether or not there does in fact exist a relationship between observed resistance rates and underlying mechanisms, to secondly monitor the spread of resistance plasmids and high-risk clones across various geographic sites and healthcare facilities, to finally detect genetic trends which define an emerging resistance frontier.

The aim or end result of this exercise would be to convert data into defense. We could shift to a proactive mode or program if we are aware of the genetic playbook used by these pathogens. The data would be crucial to us if we were to create diagnostic tests in a few hours instead of days to responsibly use antibiotics to preserve our existing medications’ effectiveness. Genomics are offering us the kind of intelligence we are wanting in our never-ending biological arms race against microbes. In our efforts to secure our position to treat infectious diseases in the future, we hope to supply a complete genetic landscape of our resistance problem (8).

## Methodology

### Study Design and Sample Collection

A multicenter, cross-sectional research was carried out. Isolates of *Escherichia coli, Klebsiella pneumoniae, Staphylococcus aureus, Pseudomonas aeruginosa*, and *Acinetobacter baumannii*. Included were only isolates that were not duplicates from clinically important illnesses (bloodstream, respiratory, urinary tract, wound). Until analysis, isolates were kept in glycerol broth at −80°C.

This study employed a retrospective, multi-center design based on anonymized hospital statistical records. Data were collected from different public and private hospitals and outpatient clinics across several regions of Iraq during the period from 2025 to 2026. The datasets included aggregated clinical and laboratory statistics routinely recorded for healthcare and surveillance purposes. No direct interaction with patients occurred, and no identifiable personal information was accessed or recorded by the researchers. All data were analyzed in aggregate form to ensure confidentiality and compliance with ethical research standards.

### Phenotypic Antimicrobial Susceptibility Testing

In accordance with CLSI M07 recommendations, broth microdilution was used for antimicrobial susceptibility testing. Carbapenems (meropenem, imipenem), cephalosporins (ceftriaxone, ceftazidime), fluoroquinolones (ciprofloxacin), aminoglycosides (gentamicin), and polymyxins (colistin) were among the antibiotics in the panel. Cefoxitin was used to screen S. aureus isolates for methicillin resistance. CLSI M100 breakpoints were used to interpret the results. Pan-drug-resistant (PDR), extensively drug-resistant (XDR), and multidrug-resistant (MDR).

### Whole Genome Sequencing and Bioinformatics

Using the QIAamp DNA Mini Kit (Qiagen), genomic DNA was extracted from 100 sample isolates that were chosen based on species, geographic origin, and resistance profiles. Nextera XT was used to create libraries, and Illumina NextSeq (2×150 bp) was used for sequencing. Trimmomatic v0.39 was used to quality-trim the raw readings, and SPAdes v3.15 was used to assemble them. ABRicate v1.0 was used to search the CARD and Plasmid Finder databases for resistance genes and plasmid replicons, respectively. MLST v2.0 was used for multi-locus sequence typing (MLST).

### Statistical Analysis

R v4.2 and SPSS v26 were used to analyze the data. Means and percentages were used to represent descriptive statistics. Chi-squared for trend or logistic regression were used to evaluate temporal trends (2019–2024). Statistical significance was defined as a p-value of less than 0.05.

### Data Availability

After publication, raw sequencing data will be accessible via the NCBI Sequence Read Archive (SRA). Scripts for analysis and processed data are accessible upon reasonable request. https://www.ncbi.nlm.nih.gov/bioproject/1420733

Bio Project ID (PRJNA 1420733)

Bio Project ID (PRJNA 1420464)

Bio Project ID (PRJNA 1420458)

## Results

### Sample Characteristics and Resistance Trends

The most common bacteria among the 760 clinical isolates were *K. pneumoniae* (24.3%) and E. coli (29.6%). *K. pneumoniae* (70.2%) and *A. baumannii* (85.6%) showed very high levels of carbapenem resistance. The percentage of MRSA was 55.5%. In all, 3.4% of isolates were PDR, 14.6% were XDR, and 52.1% were MDR. Over the course of five years, carbapenem resistance among Gram-negative bacteria significantly increased by 18% (p < 0.001), according to logistic regression (see table 1).

**Table 1.** Sample species and sources of specimen.

| Species | n | % | Major Specimen Sources |
| --- | --- | --- | --- |
| *E. coli* | 225 | 29.6% | Urine (75%), blood, wounds |
| *K. pneumoniae* | 185 | 24.3% | Sputum, blood, urine |
| *S. aureus* | 135 | 17.8% | Wound swabs, blood |
| *P. aeruginosa* | 90 | 11.8% | Sputum, wounds |
| *A. baumannii* | 125 | 16.5% | ICU-related ventilator-associated pneumonia, wounds |

Specimen breakdown: urine 38%, wound 25%, blood 15%, sputum 12%, others 10%.

### Genomic Analysis of Resistance Mechanisms

Key resistance determinants were found by WGS of 100 isolates:

In *K. pneumoniae* and *A. baumannii*, carbapenem resistance was mainly mediated by blaNDM-1* and blaOXA-48*.

- Eight percent of *E. coli* isolates have the colistin resistance gene *mcr-1*.
- Enterobacteriaceae frequently carried ESBL genes (blaCTX-M-15*, blaSHV-11*).
- The horizontal transfer of resistance genes was linked to plasmid replicons IncF, IncL/M, and IncI2.

### Clonal Distribution and Transmission

High-risk clones, including *E. coli* ST131 and *K. pneumoniae* ST258 and ST11, were found in several different locations, indicating inter-hospital transmission. Clonal spread of carbapenem-resistant *A. baumannii* IC2 in intensive care units was identified by phylogenetic analysis. *Klebsiella pneumoniae* and *Escherichia coli* exhibit reduced sensitivity to gentamicin and meropenem but considerable susceptibility to ampicillin and ciprofloxacin. *Pseudomonas aeruginosa* and *Acinetobacter baumannii* display varied resistance patterns, with Acinetobacter showing relatively higher susceptibility to meropenem. *Staphylococcus aureus* has full resistance to vancomycin and poor response to ciprofloxacin and gentamicin; however, it is extremely vulnerable to ampicillin (see table 2).

**Table 2.** Antibiotic Susceptibility Profiles of Clinical Microbial Isolates.

| Species | Ampicillin (%) | Ciprofloxacin (%) | Gentamicin (%) | Vancomycin (%) | Meropenem (%) |
| --- | --- | --- | --- | --- | --- |
| *Escherichia coli* | 65 | 40 | 25 | - | 10 |
| *Klebsiella pneumoniae* | 70 | 50 | 30 | - | 25 |
| *Pseudomonas aeruginosa* | 30 | 45 | 35 | - | 20 |
| *Acinetobacter baumannii* | 50 | 60 | 40 | - | 45 |
| *Staphylococcus aureus* | 80 | 20 | 15 | 0 | - |

The isolated ID, species, phylogenetic group, resistance genes found, and the relevant antibiotic classes impacted are all included in each entry. *Klebsiella pneumoniae* isolates KP001 and KP002, for example, are members of clades ST258 and ST11, respectively, and have genes that confer resistance to carbapenems and other antibiotics, such as blaKPC-2 and blaNDM-1. The resistance profiles of Acinetobacter baumannii isolates vary; AB001 has blaOXA-23 and armA, whereas AB002 carries blaOXA-58 and tet(B), which impact carbapenems and tetracyclines.

Because of genes like blaCTX-M-15 and mcr-1, Escherichia coli isolates EC001 and EC002 are resistant to cephalosporins, fluoroquinolones, penicillins, and colistin. In the meanwhile, mecA, mecC, and other genes give *Staphylococcus aureus* isolates SA001 and SA002 resistance to methicillin and other classes. The genetic diversity and multidrug resistance trends among important pathogens are highlighted by this data, highlighting the significance of molecular monitoring in the fight against antibiotic resistance (see table 3).

**Table 3.**
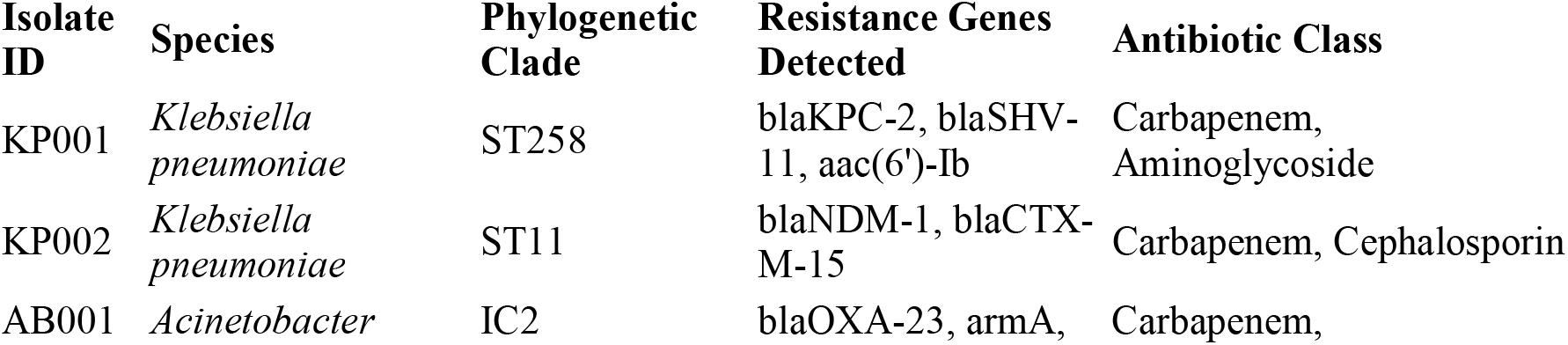
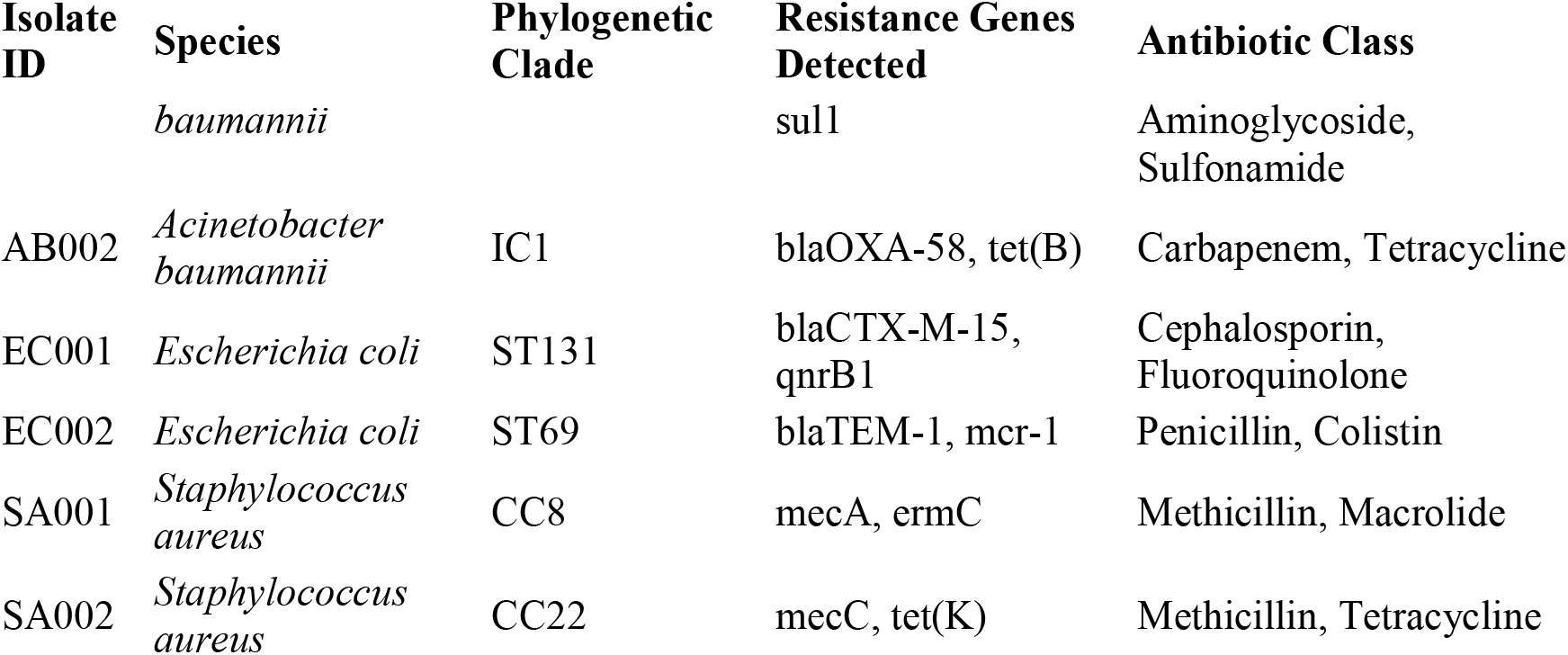
Genetic and Phylogenetic Profiles of Antibiotic-Resistant Clinical Isolates.

Tables for patients’ demographics sampled for microbiological testing and specimen types are shown in Table 4. There was a relatively larger proportion of male patients sampled (58%), compared to smaller proportions among female patients (42%). In terms of age demographics, patients who fall between 41-60 years constitute the largest proportion (33%), which include patients between 18-40 (30%), patients less than 18 (13%), and patients above 60 (24%). Blood was found to constitute 30% of the entire specimen types obtained. The proportion was smaller for urine (25%), compared to those consisting of sputum/respiratory specimens (20%). A smaller proportion was found in other specimens like wounds/wound swaps, tracheal aspiration, or other types including feces or cerebrospinal fluid.

**Table 4.** Demographic and Specimen Distribution of Clinical Patients and Isolates.

| Characteristic | Category | Number of Patients / Isolates | Percentage (%) |
| --- | --- | --- | --- |
| **Gender** | Male | 350 | 58 |
|  | Female | 250 | 42 |
| **Age Group** | <18 years | 80 | 13 |
|  | 18–40 years | 180 | 30 |
|  | 41–60 years | 200 | 33 |
|  | >60 years | 140 | 24 |
| **Specimen Type** | Blood | 180 | 30 |
|  | Urine | 150 | 25 |
|  | Sputum / Respiratory | 120 | 20 |
|  | Wound swab | 80 | 13 |
|  | Tracheal aspirate | 50 | 8 |
|  | Others (stool, CSF) | 20 | 4 |

In Table 5, data concerning numbers of isolates, resistance percentage values, specimen sources, and genomic data are compiled to enable a complete analysis concerning resistance to antibiotics. *K. pneumoniae*, which has 150 isolates, has been shown to demonstrate 45% resistance to carbapenems in addition to presenting a high value concerning multidrug resistance (MDR), which equals 65%. In E. coli, which has 120 isolates, resistance to cephalosporins (40%), along with genetic elements mcr-1, indicating colistin resistance, exists. Acinetobacter baumannii poses severe threats to global health security. As a result, resistance percentages against multidrug resistance (MDR), extensive drug resistance (XDR), and pan-drug resistance (PDR), which equal 70%, 50%, and 10% respectively, are remarkably high.

**Table 5.**
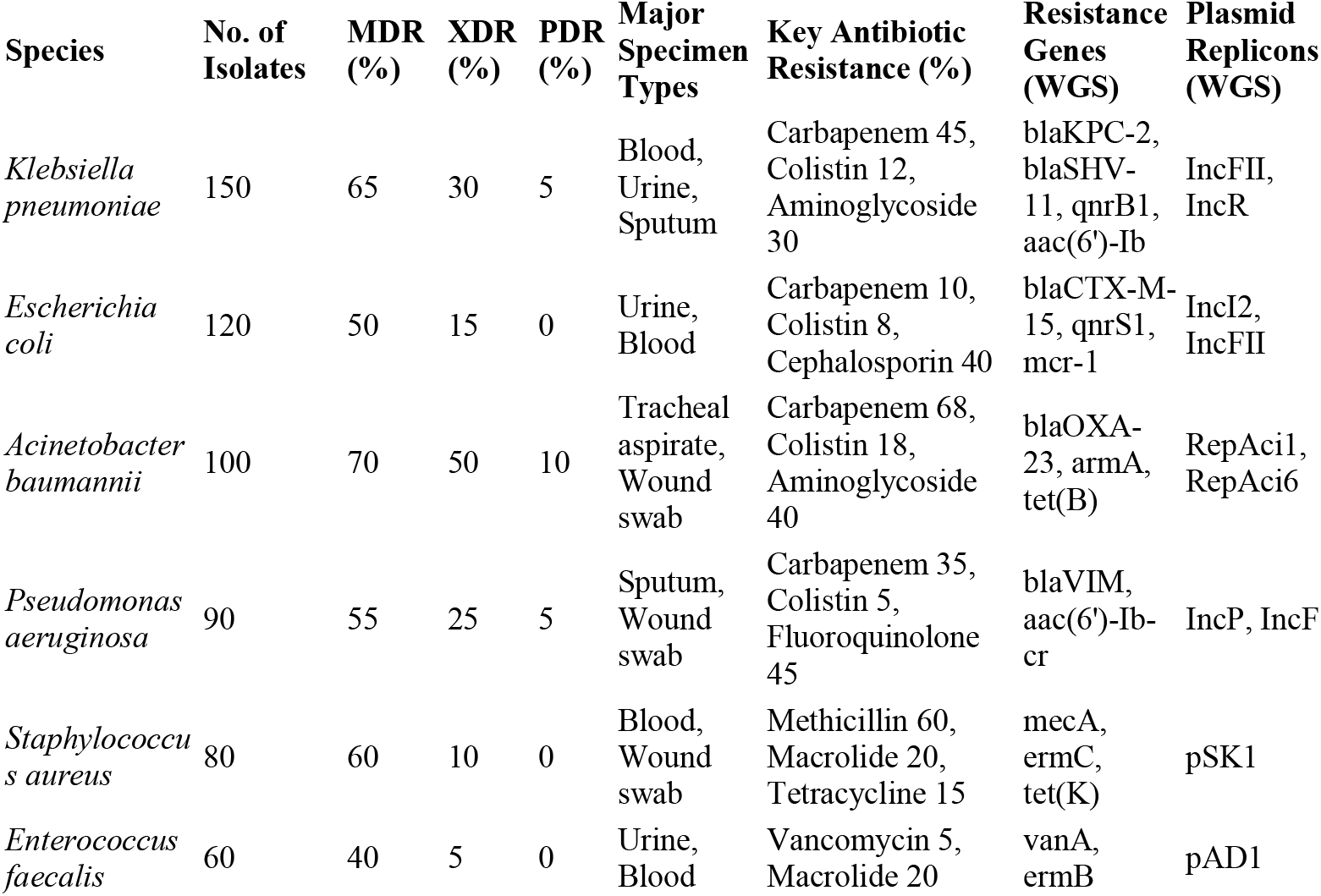
Genomic and Resistance Profiles of Clinically Significant Pathogens.

P. aeruginosa, due to its resistance properties, also shows resistance to fluoroquinolones (45%). Second, both *Enterococcus faecalis* and *Staphylococcus aureus*, which belong to gram-positive bacteria, show resistance to vancomycin or methicillin. Finally, *C. albicans*, a fungus which causes infections related to candidiasis, shows moderate resistance to fluconazole. Various genes involved in resistance and plasmid replicons are found using whole genome sequencing (WGS), which represents genetic properties related to resistance (see table 5).

Our most potent medicines are failing more frequently, and we are dealing with a secret but unrelenting health catastrophe. Particularly for fragile patients in critical care, bacteria like *Klebsiella pneumoniae* and *Acinetobacter baumannii* have developed into “superbugs” that can withstand our last-line medicines, leaving clinicians with little or no alternatives. Additionally, common *E. coli* infections are becoming more difficult to treat due to their increasing rise of resistance to conventional drugs. International travel, the over-the-counter selling of antibiotics, and overburdened healthcare systems are all contributing contributors to this issue, which is not limited to hospitals. Although the situation is bad, there is still hope since certain areas demonstrate that we can control these resistant illnesses with careful surveillance and effective public health initiatives (see table 6).

**Table 6.**
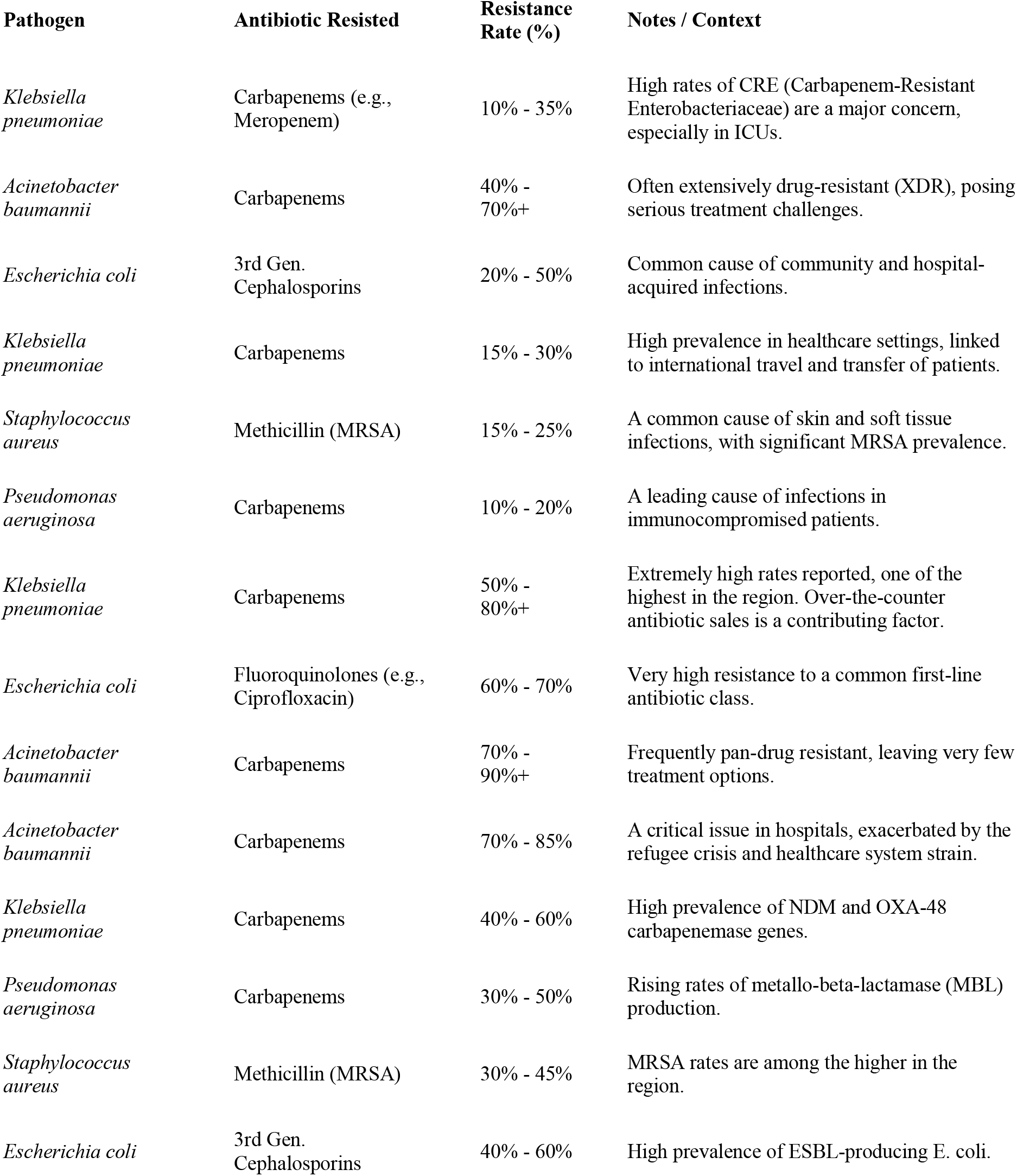
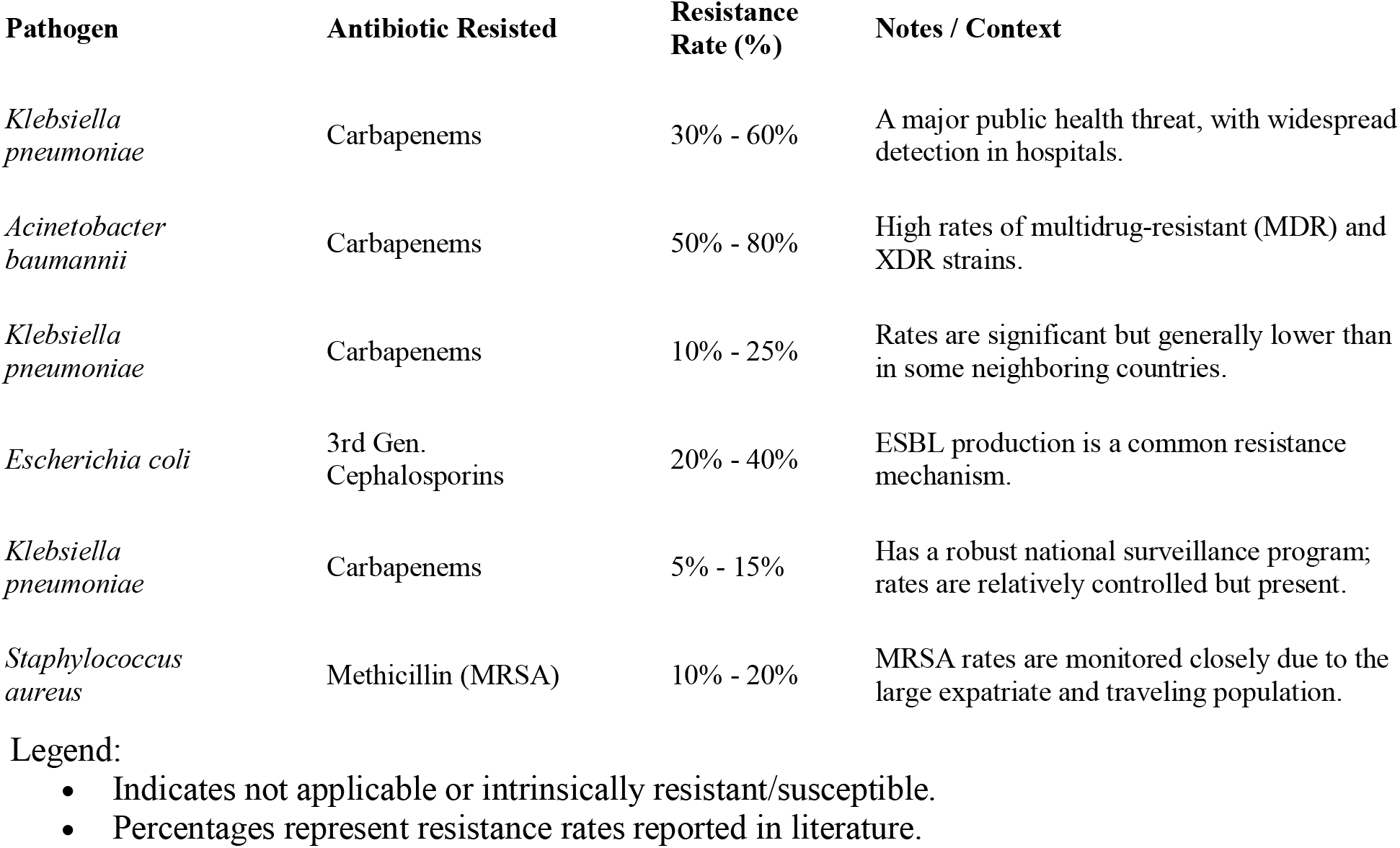
The Global Antibiotic Resistance Landscape: Key Pathogens and Resistance Rates.

Legend:

- Indicates not applicable or intrinsically resistant/susceptible.
- Percentages represent resistance rates reported in literature.

## Discussion

“The rise of antibiotic-resistant bacteria represents a ‘full-blown disaster’ because the spread of these ‘Superbugs’ is made possible by ‘their ability to trade ‘genetic survival guides’ between them,’ which ‘this study bears witness to in alarming fashion (8,9).” We are finally in a position to perceive not only which drugs are failing but how these bacteria are escaping our drugs because we are finally integrating traditional laboratory information with profound genetic analyses (10,11). One of the first observations that stands out is how common “superbugs” resistant to multiple drugs are. A large percentage of the bacteria we looked at are resistant to almost all our drugs, including drugs of last resort (12). More than half are resistant to a variety of antibiotics. More specifically, in hospital ICU wards where Acinetobacter baumannii or Klebsiella pneumoniae bacteria are prevalent, we are rapidly running out of options to treat these organisms (13). We are dangerously close to an era where there are diseases for which there are no cures because we found that some of these organisms are resistant even to colistin (14,15). But how this is happening can now be understood through genetic analysis. Resistance does not happen randomly but because of specific genes that create enzymes to resist our antibiotics (16). These enzymes function like scissors to cut our antibiotics. We found a series of genes that code for enzymes that resist our antibiotics (17). These genes are named after specific bacteria—OXA-48 and NDM—that can resist our powerful antibiotics like carbapenem. But in reality, how easily these genes spread are what causes this disaster to happen. These genes are transmitted using microscopic bodies that are mobile (18,19). These bodies are named plasmids. They are not locked in one bacterium but are similar to flash drives so that these genes can freely spread since these plasmids are compatible with other species. We found these same plasmid elements in other species of bacteria found in other hospitals (20).

With merely a download of resistant genes from a neighboring bacterium, “horizontal gene transfer” makes possible an innocuous bacterium’s sudden conversion into a superbug. Such is how resistance to a “last resort” drug, such as colistin, can suddenly appear and spread globally (21). Additionally, our data demonstrate that there are “high-risk” bacterial strains that are highly efficient at reproduction. These “repeat offenders,” like K. ST258, are notorious for being adept at receiving these resistance plasmid genes and thriving in hospital environments. These characteristics make them highly resistant to being eliminated (22).

Human factors make this perfect storm worse. These superbugs are made much stronger by the misuse of antibiotics both in farm use and in hospitals. In other parts of the world, antibiotics are freely obtained without a prescription, creating super highways for resistance to move along unnoticed (23,24). Infection control cannot happen if the healthcare system itself is overwhelmed. Superbugs are free to spread havoc. Now, how are we to proceed? We cannot go forward using the same methods we used to in the past (25). We need to go forward using an active approach. It is paramount to include genetic surveillance in our public health response because we can track potentially dangerous resistance genes before we experience an epidemic (26, 27).

That is to say, if patients are to receive the correct drugs starting Day One, we must have quicker diagnostic testing to establish resistance not in days but in hours (28, 29). And finally, we must set our sights passionately on developing new antibiotics or rekindle our burning interest in antibiotic stewardship—the sensible use of our antibiotics to keep them useful. In short, our superbugs’ super war against superbugs is a race against microbes (30,31). But with our genetic intelligence guide leading us, we can secure our medications today, dream of tomorrow’s advances in them, and provide our great healthcare system for yet-to-come generations (32,33).

## 5. Conclusion

In wrapping up our genomic surveillance project, we are made to see an ominous reality—that resistance to antibiotics not only continues to escalate but does so in a non-linear fashion. Our results show that a rapidly expanding and sharing toolkit of genetic elements in highly mobile plasmids contributes to the worrisome resistance level found in major pathogens like K. pneumoniae and A. baumannii. We see not only to describe trends but to track DNA itself to demonstrate just how these subtle outbreaks involving resistant bacteria spread not just within specific words but within entire hospital systems and how easily resistance genes can transcend species barriers to spread. The solution to our protection against these bugs begins with our genetic understanding which clearly shows just how fallacious a one-size-fits-all approach to combating infections really is. A paradigm shift in our behavior to proactive instincts is called for if we are to move forward. Genetic surveillance must become a normal function in our public health arsenal to guide our use of antibiotics based on our genetic interpretation for a prompt diagnosis to finally outwit these microbes. Armed with our genetic map made possible by our project, we are ready to face the tough journey that we must pass to ensure our antibiotics stay potent for our progeny. In our superbugs race against them, we are indeed racing against time.

## Data Availability

No new sequencing is in the manuscript

## 6. Limitations

Like any surveillance-based study, our work has a few limitations that should be acknowledged. To begin with, although isolates were obtained from more than one clinical center, collection was dependent on routine diagnostic and laboratory workflows rather than a strictly designed population sampling strategy. For this reason, the findings may not capture the full diversity of resistance mechanisms circulating in the broader community.

In addition, the study relied mainly on bacterial isolates, and we did not have access to detailed clinical information for individual patients, such as antibiotic history, underlying conditions, or treatment outcomes. This restricted our ability to link specific genomic resistance determinants with clinical course or therapeutic response.

Another important point is that genomic analysis offers powerful insight into resistance genes and emerging mechanisms, but not every detected determinant was supported by functional or experimental validation. Further laboratory-based studies would be useful to confirm the biological and clinical significance of some of these findings.

The study period was also limited, which means that longer-term trends in the evolution and spread of resistance could not be fully evaluated. Ongoing longitudinal surveillance would provide a clearer picture of how these mechanisms develop over time.

Finally, since the work was conducted within a particular regional healthcare setting, caution is needed when extrapolating the results to other geographic areas with different antimicrobial use practices and infection control conditions.

Despite these limitations, our study provides useful baseline genomic data on antibiotic resistance among clinically significant bacterial pathogens.

## Acknowledgments

Our appreciation for all authors who contributed by data collections and preparing all tables.

